# Do panic values cause panic? Reporting of critical laboratory results in a tertiary hospital in Kenya

**DOI:** 10.1101/2022.04.25.22274278

**Authors:** Thomas Mwogi, Tim Mercer, Dan N. (Tina) Tran, Ronald Tonui, Thorkild Tylleskar, Martin C. Were

## Abstract

Prompt communication of critical laboratory results is important for patient safety. Various standardisation bodies have proposed procedures for handling critical results, with notification parameters outlined. However, few studies exist in low- and middle-income countries (LMIC) to document how critical results are handled. We tracked 12 types of laboratory tests over a three-week period in December 2018 and documented if and how critical test results were communicated, the time-frame for communication, and evidence of action taken on the results. During the period, 331 of 5,500 (6.1%) test results were identified as critical. Only 71 (21%) of the critical results were documented as having been communicated to the destination departments. Of the communicated results, clinicians were unaware of 21 (29.6%). Of the 12 test types, critical results were only communicated for three tests namely: potassium, haemoglobin and positive malaria tests. Communication of critical results to inpatient settings was significantly higher than to outpatient settings (p <0.05), with communication rates decreasing as the week progressed, during weekends and around holidays. The observed poor communication of critical results in an LMIC setting raise significant patient safety concerns. Laboratories in these settings need to adhere to international standards, like ISO 15189:2009, to assure safe practice. Training of staff, establishment of standard operating procedures guiding these results, and implementation of fail-proof critical result dissemination mechanisms are essential. It is important that all critical results are communicated within one hour of availability. Implementation of Order Entry and Laboratory systems should be highly considered.

## Introduction

Critical laboratory values also known as panic values, were described by Dr. George Lundberg more than 30 years ago as laboratory values that suggest that the patient is in imminent danger unless prompt and appropriate action is taken to avert it [1]. A critical value represents a pathophysiological state so far away from the normal value to potentially be life threatening unless a timely corrective action is taken. The Joint Commission, an independent and not-for-profit organization in the United States defines a critical result as a ”test that requires immediate communication of result irrespective of whether it is normal, significantly abnormal or critical” [2].

Communication of a critical test result involves the relay of this results to the clinician or the nurse taking care of the patient for necessary action to avert further harm to the patient. To differentiate critical results from other results, some authors have introduced the concept of *’must know now*’ to describe critical results [3].

The importance of critical values became official when the concept of panic or critical values was incorporated into the Clinical Laboratory Improvement Amendments in the United States [4]. Subsequently, the requirement of having the caregiver write down and read back the critical value information (so-called read-back) was introduced as a means of improving patient safety [5, 6]. The laboratory typically receives patient samples, runs the tests, and relays the results back for action by the clinical team. How critical a result is determines the expected speed of this communication. No universally agreed standard for determining which tests should be in a hospital critical list exists. A Q-Probes study of 163 laboratories determined that multiple methods are used by institutions in determining which tests should be regarded as critical [7]. About one third of the laboratories used published literature sources, another third used non-laboratory medical staff recommendations, and one third used other sources such as internal studies, inter-laboratory comparisons, or manufacturers’ recommendations.

Tillman and Barth [8] conducted a survey of hospital biochemistry laboratories in the UK. Of the 94 laboratories, 23 obtained concurrence on the alert limits with their doctors, experience and the literature. Two laboratories quoted literature to support their values, while seven laboratories did not submit actual panic values. There was discrepancy in the values interpreted as critical by the laboratories. The study team recommended that every laboratory needs to appraise its list of panic values while aiming for a modest number of analytes that are always conveyed to the clinicians, with sensitivity not to overburden the providers.

In another instance, Arbiol-Roca et al [9] performed an analysis of critical values on data obtained over a six-month period in a tertiary university hospital in Spain. Of the 5,723 alert values, 4,577 (80%) came from point-of-care testing, 884 (15%) from routine inpatients testing, and 262 (5%) from routine outpatients testing. The dominant portion of panic values was oxygen partial pressure (17.7%), followed by potassium ion concentrations (17.6%). Inpatients’ parameters reported as critical were sodium ion, phosphate, haemoglobin, glucose and potassium ion concentrations while among outpatients it was potassium and calcium concentrations.

The College of American Pathologists and others have made recommendations for improving how critical tests should be handled within institutions [10]. While the set and threshold of critical values might differ from one institution to the other, every institution needs to define its criteria of what is deemed critical for all relevant laboratory tests [7]. Once an institution determines their list of critical tests and critical values, the next logical step is to design and implement a standard operating procedure (SOP) for communicating these results. Such an SOP was published by the ‘Massachusetts Coalition for the Prevention of Medical Errors’ to improve communication, teamwork and information transfer for critical values [11] The SOP addresses the following questions: (1) Who should receive the results, (2) Who should receive the results when the ordering provider is not available, (3) What results require timely and reliable communication, (4) When the results should be actively reported to the ordering provider with explicit time frames, (5) How to notify the responsible provider and (6) How to design, support, and maintain the systems involved.

Timely reporting of critical values is an accreditation requirement for clinical laboratories in most countries. The ISO 15189:2012 is widely adopted by a large number of laboratories outline in detail how critical values are to be handled [12]. Sub-clause 5.5.3 (q) of the standard requires that examination procedures should be documented when applicable to the examination procedure to include critical values. Sub-clause 5.9.1 (b) of the same standard prescribes that for critical results, the medical laboratory should establish procedures for immediate notification of the physician or other authorized health professional. It also recommends that the notification attempts, the results conveyed and any difficulties encountered during the notifications should be documented. In general, critical results and values are crucial to patient safety. It is the expectation that they be acted upon promptly to avert loss of life.

While guidelines for handling critical test results are widely available, hardly any studies have been done in low and middle-income countries (LMIC) to document how such critical results are actually managed. A study in Tygerberg Hospital in South Africa audited the accuracy of telephone communication of critical results and found a 10.8% error rate for accuracy between the critical results and what was communicated and recorded at the destination [13].

## Methods and methods

### Setting

This was a retrospective, descriptive, study of selected critical laboratory tests at a tertiary hospital in Kenya conducted in 2018. The hospital has achieved ISO accreditation in Quality Management Systems (ISO 9001:2015 Standard) and was also a certified Medical Laboratory Standard (ISO 15189:2012 Standard) hospital. At the time, the hospital had 11 clinical laboratories, namely: hematology, biochemistry, microbiology, tuberculosis, immunology, histology / pathology, parasitology, blood bank, blood transfusion unit, private wing and children’s unit laboratory. The laboratories operated 24/7 and testing was run continuously although samples are received in batches. The hospital laboratories handled a volume of a million tests per year, and the laboratories were staffed by 174 officers distributed between the different laboratories. These laboratories processed all samples from every inpatient and outpatient setting of the hospital, with no tests sent to outside laboratories. The inpatient wards were staffed by phlebotomy officers who picked up paper requests from clinicians, took samples from patients and delivered these samples for processing in the relevant laboratory. Generally, there was a close and harmonious working relationship between the laboratory personnel and the practitioners.

The laboratory has a standardized procedure for communicating critical results. The procedure stipulates the confirmation and review of a new critical result. The critical result then needed to be promptly communicated via a telephone call to the clinican or the nurse taking care of the patient. The communication was to be documented in a critical value reporting register.

### Patient/Public Involvement

Patients and the public were not involved in the design, conduct, reporting or dissemination of this work as they were not directly involved at any of the points.

### Study question

The aim of this study was to answer several questions around critical results at the hospital, namely: (1) What was frequency with which critical laboratory results occurred, (2) what was the distribution of critical laboratory results among the different types of laboratory tests, (3) were the results communicated to their destinations as per the institutional SOP, (4) what mode was used to communicate critical results, (5) were clinians aware of the result, and (6) what action was taken based on the critical result.

### Tracking panic values

For this study, we tracked the workflow for 12 laboratory tests and associated critical values (Table 1) based on well-accepted criteria for critical tests outlined by the College of American Pathologists [10]. Data were collected over a period of three weeks in the month of December 2018. This period was selected because the last two weeks of December are a holiday period for staff at the hospital. It was deemed likely there would be delays during this period not identified by past studies.

**Table 1.** The tests and critical results tracked in the study.

| Test # | Critical values criteria |
| --- | --- |
| 1 | Potassium less than 3mmol or above 6mmol/l |
| 2 | Sodium greater than 170mmol/l or less than 110mmol/l |
| 3 | Creatinine more than 600 mmol/l |
| 4 | Chloride less than 75 mmol/l |
| 5 | Troponin more than 0.1 mg/l or 4 pg/ml |
| 6 | Haemoglobin less than 6.6 g/dl or greater than 19.9 g/dl |
| 7 | Leukocyte count less than $2 \times 10^9/l$ or greater than $5 \times 10^9/l$ |
| 8 | Platelet count less than $20 \times 10^9/l$ or greater than $1000 \times 10^9/l$ |
| 9 | Malaria Positive |
| 10 | Peripheral blood film Sickling Test Positive |
| 11 | Blood culture Positive |
| 12 | INR greater or equals 5 |

Every day during the 3-week study period, trained research assistants (RA) would scan through the previous day’s results in the laboratory results register. Results meeting the criteria of critical results (Table 1) were included in the study. The RA would record the patient demographics, time of result availability to clinicians, destination, the test type and the value of the test result. A communications’ register in the laboratory was used as source of information for whether the results had been communicated. If present, the time of communication, type of communication and the person communicated to were recorded. If the communication was missing, it was deemed to not have been communicated.

The RA also checked for evidence of communication at the destination. For this, the RA would check the nursing documentation, the patient chart, ward or clinic register and as feasible through verbal confirmation with care providers in the destination department for evidence of communication. In addition, the RA would also check the patient chart, prescriptions, medication sheet or the nursing notes for evidence of action on each critical test result. If evidence of action was present, the time of action as well as any action taken, were also recorded. A majority of the actions taken were related to lowering to blood potassium levels for hyperkalemia, initiation of antimalarials and blood transfusion for malaria and anemia respectively. All findings were recorded in the secure REDCap application and stored centrally in a secured database [14].

### Ethical Considerations

The study was approved by the Institutional Review and Ethics Committee at Moi University College of Health Sciences/Moi Teaching and Referral Hospital and the need for consent was waived having been regarded as a quality improvement research with anonymization of data. The project was also assessed by the Regional Committee for Medical and Health Research Ethics in Norway (2017/2501/REK vest) as a quality assurance project and therefore outside REKs remit.

### Data analysis

The collected data were extracted by the study team member (TM) from the REDCap database and patient identifying information were removed [14]. Study personnel scanned these data for any inconsistencies in timestamps and data recorded, missing or invalid data. Quantification of all tracked results was done and percentages calculated for communicated and non-communicated results. Results were also classified based on destination. The mean, median and standard deviation of the time differences between result availability and result communication were calculated. In order to assess any relationship between the test type, day of the week and communication of the results, the T-test and chi-square tests were used.

## Results

### Overall

A total of 331 critical test results out of 5,500 (6.0%) were identified over the three week study period. Of the critical results, 71 (21.5%) were documented within the laboratory results register at the lab as having been communicated to the destination unit while 260 (78.5%) were not communicated. The communicated results were only for a few tests, namely: potassium (both critically high or low levels), hemoglobin (critically low levels only), low leukocyte count, and positive malaria test result (Table 2). None of the critical results for the other tests were communicated (Table 1). Communicating of critical results was done solely through phone calls to either a receiving nurse or doctor in the destination departments.

**Table 2.** Proportion of critical results communicated. *The standard is for 100% of critical results to be communicated directly to providers from the lab. .

|  | Critical tests | Total | Communicated | % | Chi-square |
| --- | --- | --- | --- | --- | --- |
| 1 | Potassium <3mmol or >6mmol/l | 75 | 63 | 84.0 | p <0.01 |
| 2 | Sodium <170mmol/l or >110mmol/l | 7 | 0 | 0.0 | p = 0.17 |
| 3 | Creatinine >600 mmol/l | 50 | 0 | 0.0 | p <0.01 |
| 4 | Chloride <75 mmol/l | 18 | 0 | 0.0 | p <0.05 |
| 5 | Troponin >0.1 mg/l | 13 | 0 | 0.0 | p = 0.06 |
| 6 | Haemoglobin <6.6 g/dl or >19.9 g/dl | 73 | 6 | 8.2 | p <0.01 |
| 7 | Leukocyte count <2000/ml or >50,000/ml | 41 | 1 | 2.4 | p <0.01 |
| 8 | Platelet count <20,000/ml or >1 million/ml | 39 | 0 | 0.0 | p <0.01 |
| 9 | Malaria test positive | 3 | 1 | 33.3 | p = 0.62 |
| 10 | PBF sickling test positive | 3 | 0 | 0.0 | p = 0.37 |
| 11 | Blood culture positive | 6 | 0 | 0.0 | p = 0.20 |
| 12 | INR >= 5 | 3 | 0 | 0.0 | p = 0.37 |

As shown in Table 3, most critical test results (224 of 331, 67.3%) originated from tests conducted in the inpatient setting, while the rest originated from the outpatient clinics (41 of 331, 12.4%) and operating rooms (66 of 331, 19.9%). The percentage of communicated results to the inpatient wards was significantly higher compared to those communicated to the outpatient departments or operating rooms (p <0.05).

**Table 3.** Frequency of critical results communication by across key hospital areas

|  | Destination of communication | Total Results | Communicated | % | Chi-square |
| --- | --- | --- | --- | --- | --- |
| 1 | Inpatient ward | 224 | 64 | 28.6 | p-value = .000578 |
| 2 | Outpatient departments | 41 | 4 | 9.8 |  |
| 3 | Operating rooms | 66 | 3 | 4.5 |  |
|  | Total | 331 | 71 | 21.5 |  |

Of the 71 critical results that were documented by the laboratory as having been communicated successfully, only 58 (81.6%) had documented evidence of this communication in the destination departments – i.e., recorded as having been communicated by the laboratory with the ward, clinic or emergency room registers. However, those with actual evidence of action were only 57 (80.2%). Clinical providers were only aware of 21 (29.6%) critical results despite action having been taken against them by other providers through orders or clinical notes. (Figure 1).

**Fig 1.**
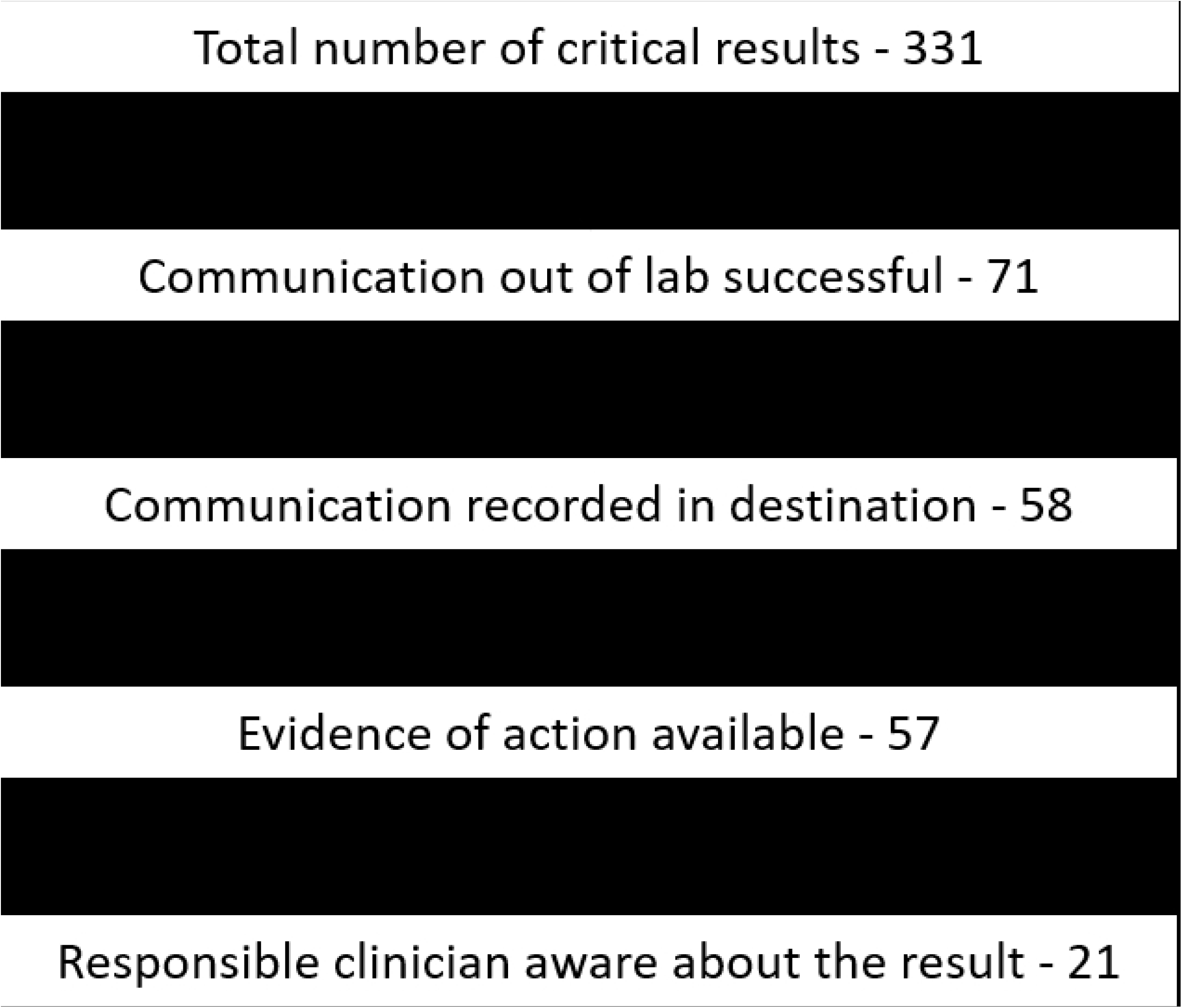
Sequence of communication. The figure shows the sequence and drop off along the chain from communication to action on critical results.

A further analysis of the critical tests was done to determine if communication was affected by the day of the week or the day of the month. Table 4 shows the relationship between the day of the week when the critical result occurred, and the percentage communicated. Results from early in the week were significantly communicated when compared with the mean (21.5% communication rate overall). There was low communication rate for Wednesday, Thursday and Friday.

**Table 4.** Table showing the relationship between the days of the week when results were communicated and the success of communication

|  | Day of the week | Total Results | Communicated | % | Chi-square |
| --- | --- | --- | --- | --- | --- |
| 1 | Sunday | 36 | 20 | 54.6 | **p < .01** |
| 2 | Monday | 36 | 14 | 38.9 | p = .01 |
| 3 | Tuesday | 65 | 13 | 20.0 | p = .79 |
| 4 | Wednesday | 51 | 6 | 11.8 | p = .11 |
| 5 | Thursday | 67 | 4 | 6.0 | **p < .01** |
| 6 | Friday | 34 | 3 | 8.8 | p = .08 |
| 7 | Saturday | 43 | 11 | 25.6 | p = .54 |
|  |  | 331 | 71 | 21.5 |  |

Figure 2 shows the relationship between the day of the week and the time it took for caregivers to act on critical results after they were available. As this data was collected in the month of December, the figure shows that it took longer to act on critical results as the holiday season approached.

**Fig 2.**
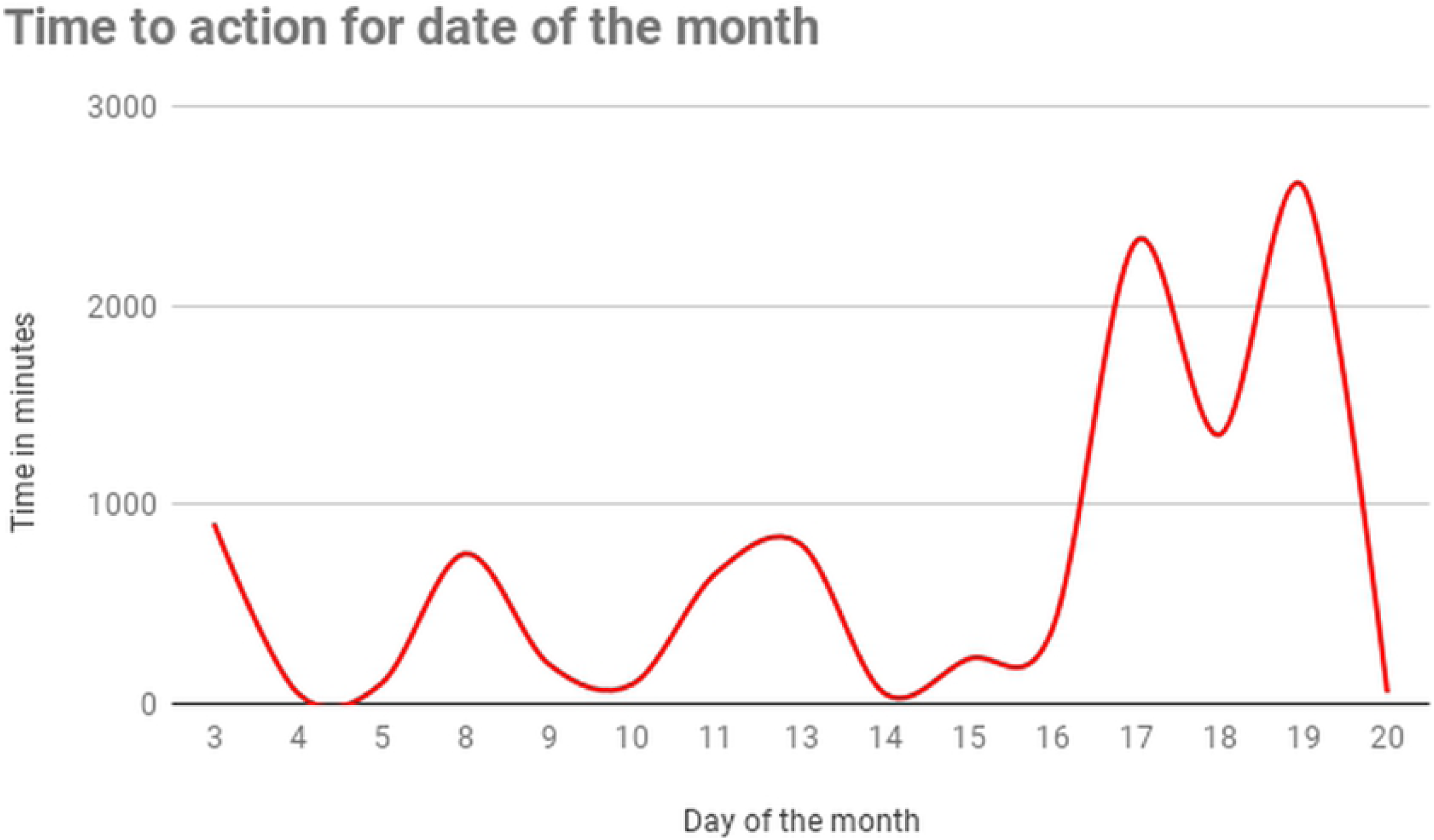
Time to action. The figure shows the average time it took to act on critical results after they were available for each day of the week.

Figure 3 shows the results communicated for the period. Most results were communicated with a median time of 45 minutes with some outlier results being acted upon after up to 1,503 minutes (25 hours) with a mean time to action of 200 minutes (SD = 362 minutes).

**Fig 3.**
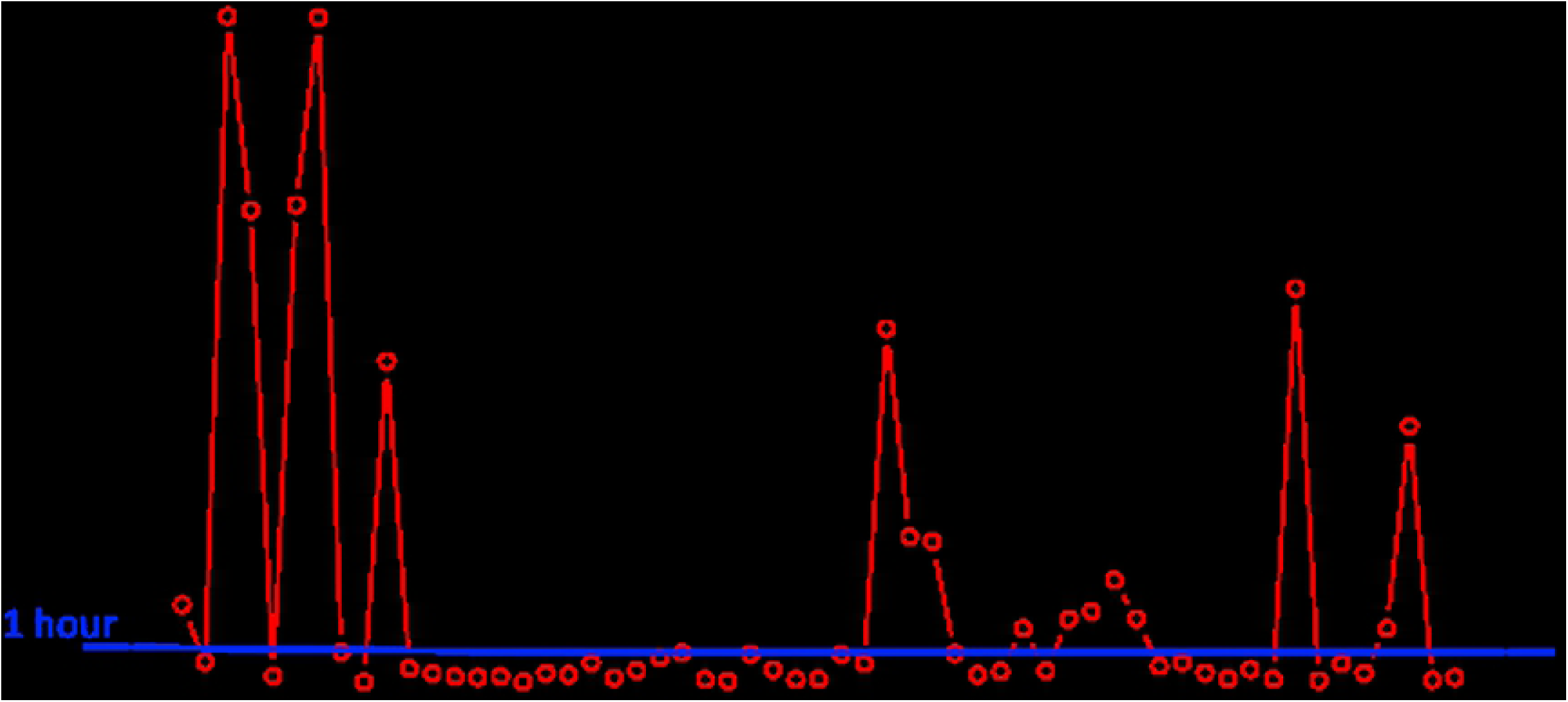
Time to action. The figure shows the average time it took to act on critical results.

## Discussion

In this study, we observed a very low rate of communication of critical laboratory results in a tertiary care setting within an LMIC setting. Overall communication rates for the critical results was only 21%, which is significantly lower compared to other studies where rates of critical result communication range from 60% to over 90% [15] likely highlighting a patient safety concern. Lack of reporting of critical results may have had a direct impact on morbidity and mortality [15, 16]. It was observed that certain critical tests were more likely to be communicated than others despite the official laboratory policy indicating importance of reporting all critical values promptly [2].

There was a high drop off rate in the process of communicating critical results. Only 58 of the 331 critical results (17.5%) were evidently communicated. It was also noted that there was no callback for successfully communicated critical results although this requirement is captured in the critical value reporting procedure [17]. It is not just important for results to be communicated. A complete communication cycle involves read back to ensure successful interpretation of the communication at the receiving end. There should be evidence of communication and evidence of action in the destination department. Read back of communicated critical values is now a universal mandatory recommendation across a variety of guidelines [16].

One hour is now almost universally agreed as a time limit for reporting critical results [16]. A majority of communicated critical results in this study were done within a median time of 45 minutes. Reporting was higher for inpatient setting when compared to outpatient setting in our study. This is consistent with other studies as well [9, 15] which found reporting of critical values in outpatient setting to be problematic. Notification systems for outpatient settings have been found to be largely inadequate when compared to inpatient settings and there is need to put more effort in improving these systems. This is especially important given that the patient may have been allowed to leave the hospital setting. There was a significant influence on the day of the week or holiday period on successful communication. It is postulated that due to high workload, laboratory staff tend to tire towards the end of the week. Appropriate staff-workload ratios have to be maintained to ensure adequate communication of critical results.

Given the impact of critical results communication breakdown on morbidity and mortality it is important that the problem is promptly sorted. Multiple solutions need to be implemented in order to tackle the problem. These solutions include: (1) Training and sensitization of staff, (2) frequent review of official laboratory standard operating procedures (SOPs) on management of critical results, (3) proper implementation of adopted standards (ISO and others), (4) addressing staff workload issues and (5) implementing technology solutions. There is need for training of laboratory staff on communication workflow, escalation for unsuccessful communication, read back and adherence to the laid down SOPs. Laboratory staff in collaboration with clinicians need to come up with a list of mutually agreed critical results suited to the conditions encountered in the hospital. There is need for the management to fully implement the standards already adopted e.g. ISO 15189:2012. Additionally, the hospital should reassess the laboratory staff needs to ensure proper staffing and balanced workload regardless of the time of the month.

The selection of critical value thresholds has been looked at in various studies [18–20]. One study extracted anonymized laboratory results of patients over eighteen years old from an intensive care database. The bottommost and uppermost critical alert thresholds were acquired from the forthcoming lowest and highest laboratory values, which correlate to anticipated chance of demise at 90%. The study concluded that the incidental approach applied was a realistic way to obtain threshold values that are clinically worthwhile [18]; on the other study, 10 laboratory tests with the strongest association with death in descending order were identified as: bicarbonate, phosphate, anion gap, white cell total count, partial thromboplastin time, platelet, total calcium, chloride, glucose and INR [19]; a systematic review of literature on alert thresholds for common biochemical and hematological tests in adults indicated from another inquiry that 70% of papers reported thresholds set by individual institutions, 18% contained thresholds from surveys of laboratories, 46% of the papers referred to 1 or both of the 2 American laboratory explorations from the beginning of 1990s [20].

A study in large tertiary hospital in Singapore reported the use of SMS to notify critical laboratory results [21]. The text messaging system allowed the physician to respond by acknowledging or rejecting the panic alert by SMS reply. An automated escalation is produced after ten minutes if there is no confirmatory receipt. The implication of that was a decrease in median time from 7.3 minutes to 2 minutes. That was definitely a very useful solution that helped to quickly report panic values to targeted clinicians for timely mediation.

Technology has been shown to improve reporting of critical results and to inform clinicians of critical events just in time [22]. Implementation of a well-designed laboratory information system integrated into computerized physician order entry (CPOE) systems could increase the percentage of reporting, introduce real-time alerts targeting care providers, escalate the alert in cases of no interventions within a certain time and reduce staff workload. CPOE systems provide the opportunity to closely tie the critical results and monitoring of action against the result. In one study, implementing CPOE systems and other measures increased critical result reporting from 55% to 95% within four years [23]. CPOE systems also offer additional advantages. They may prevent serious medication errors, adverse drug reactions, drug-drug interactions, and may improve physician performance and overall patient outcomes [22, 24, 25]. They have also been shown to directly influence the time to action [26].

Paper-based test requisition forms additionally come with disadvantages. Ineligible writing often make it difficult for laboratory staff to discern important clinical information that could have informed on urgency. The ineligibility may also prevent staff from determining who the responsible clinician is [15]. In our previous paper, we found that paper based requisition and results delivery processes contributed to significant delays [27].

There are limitations of this study. The study was quantitative in nature. A further qualitative component to explore the complex relationship between clinicians, phlebotomists, laboratory staff and the communication cascade may be needed in order to fully understand potential undocumented bottlenecks. This study did not collect any data on clinical outcomes and therefore did not link the reporting (or lack) of critical results to outcomes. It was also conducted in a single large tertiary hospital and the results may not be generalizable to other hospital settings in low- and middle-income countries. Furthermore, we relied on documented data. Some results that may have been communicated but not documented and therefore not included in this study. We did not include point-of-care (POC) tests, e.g., blood sugar.

## Conclusion

In low resource settings, multiple challenges often affect optimal provision of care to patients. Laboratories may suffer from lack of proper equipment and reagents, inadequate personnel for the workload, erratic power supply among other challenges. Critical results despite being very essential to patient care may be overlooked. There is need to fully adopt recommendations across different guidelines including the ISO 15189:2012. It is important that every effort is made to ensure all critical results are communicated within one hour of availability and that all communication is read back for validation. Implementation of Clinical Provider Order Entry and Laboratory Information Management systems may go a long way in solving these issues as well as reducing the workload in settings where employment of more staff may not be an alternative.

## Data Availability

The data underlying the results presented in the study are available from the Author or through access to the REDCap database (https://research.kenyamedicine.info) - registration is free and approval to the project will be done through a request on the home page after registration.

## Acknowledgments

This work was supported in part by the NORHED-funded project Health Informatics Training and Research in East Africa for Improved Health Care (HI-TRAIN, QZA-0484). Partial funding was also received from Moi Teaching Referral Hospital as part of its quality improvement initiatives and research support. The content is solely the responsibility of the authors and does not necessarily represent the official views of the Norwegian Agency for Development Cooperation or of Moi Teaching Referral Hospital.

We acknowledge the immense support received from the hospital management and more specifically by the Chief Executive Officer, the head of the department of laboratory services Ms. Florence Tum and the deputy of the same department Mr. Philemon Chebii. We thank the research assistants: Carolyne Songok, Millicent Tanui and Olympia Cheruiyot for their dedication and attention to detail as well as going beyond their call of duty to ensure work done was as perfect as humanly possible.

## Appendix 1 Instruments Used

**Fig 4.**
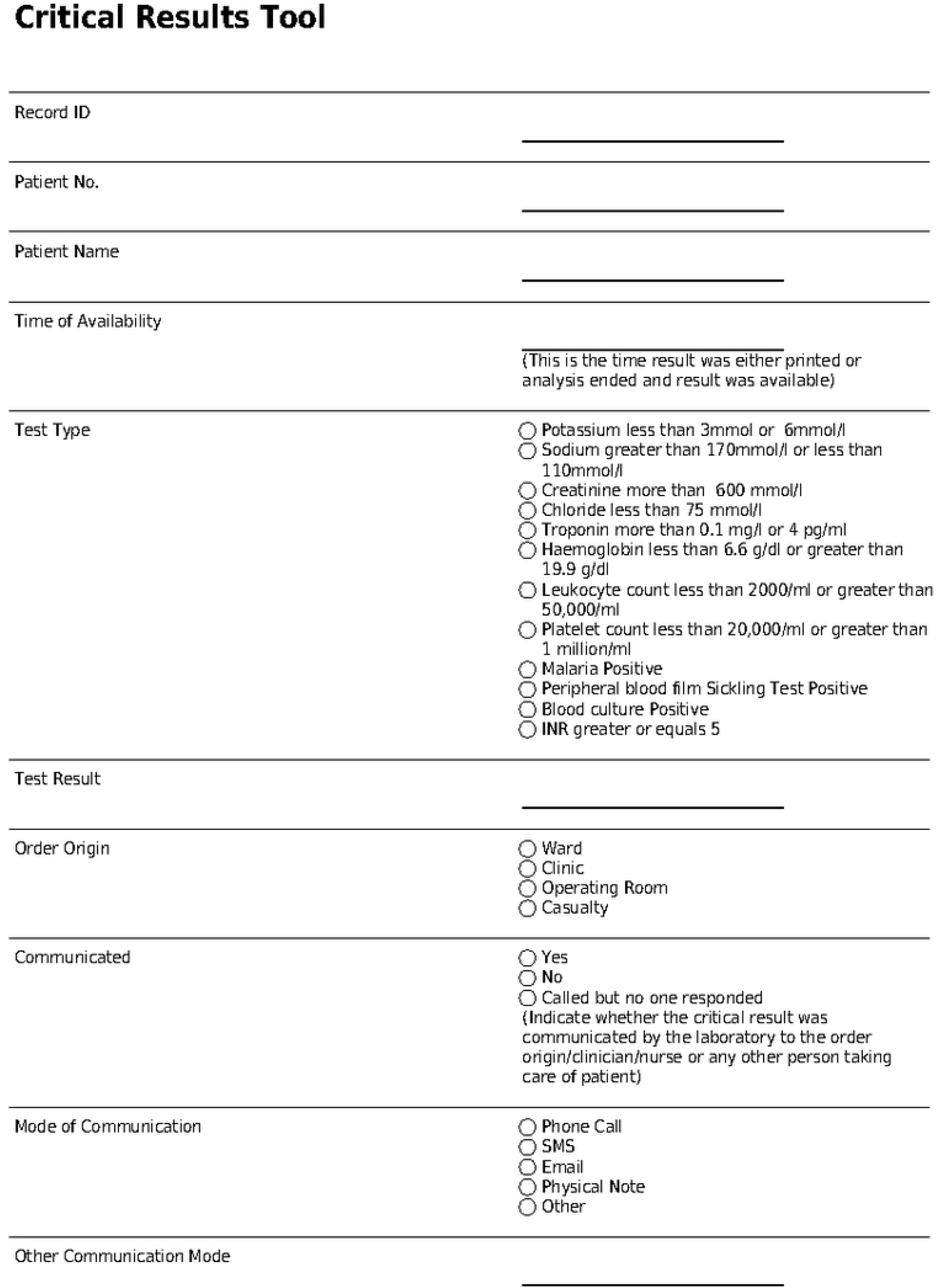
Critical results tool. Part 1.

**Fig 5.**
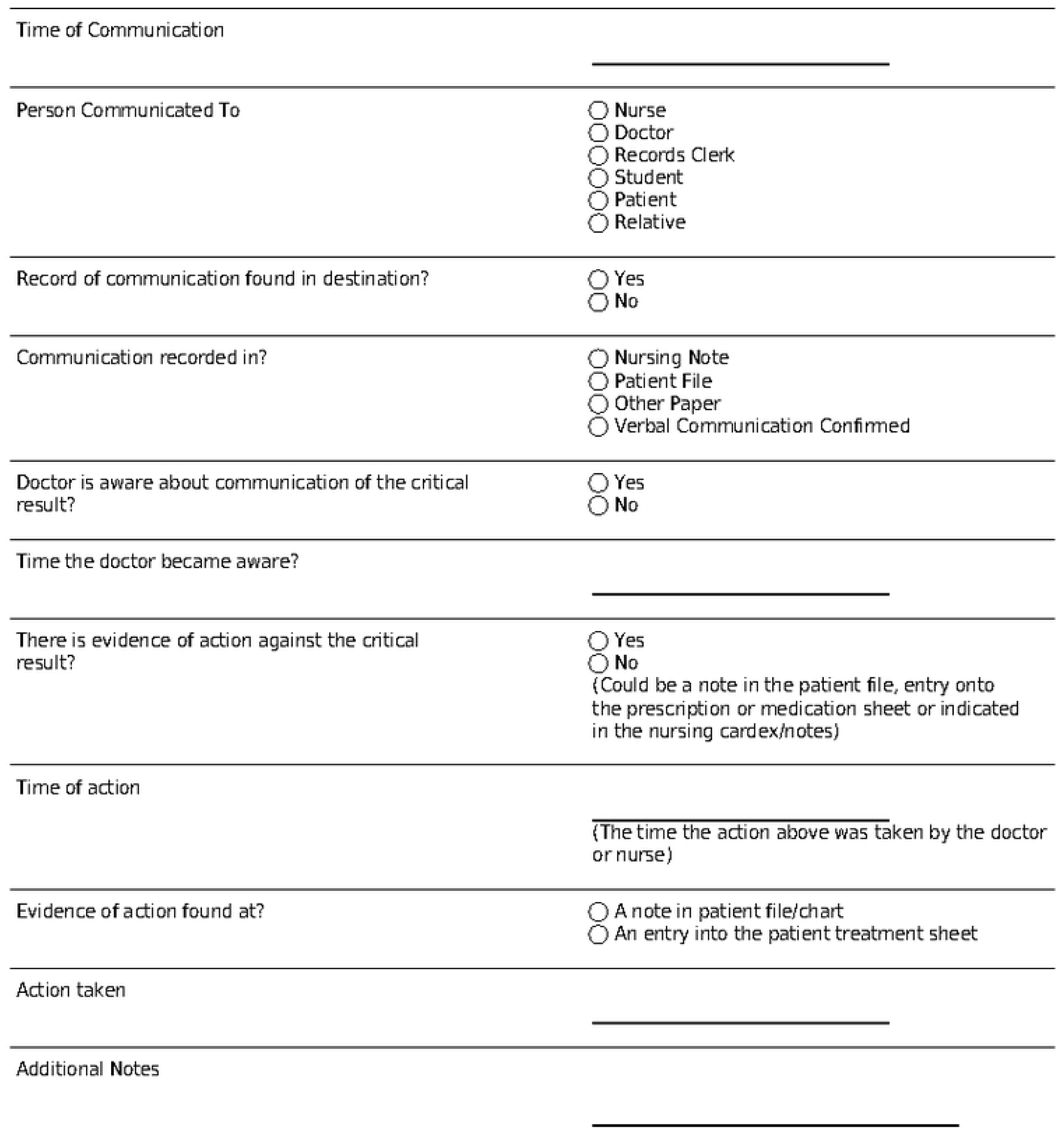
Critical results tool: Part 2.

